# Simple and economical RNA extraction and storage packets for viral detection from serum or plasma

**DOI:** 10.1101/2022.01.28.22270041

**Authors:** Sarah Hernandez, Fátima Cardozo, David R. Myers, Alejandra Rojas, Jesse J. Waggoner

**Author notes:** Corresponding Author: Jesse J. Waggoner, 1760 Haygood Drive NE, Room E-169, Bay E-1, Atlanta, GA, 30329.

## Abstract

RNA extraction is an essential step for detection and surveillance of common viral pathogens. Currently, sample processing and RNA extraction are costly and rely on proprietary materials that are difficult to acquire, maintain, and safely discard in low-resource settings. We developed an economical RNA extraction and storage protocol that eliminates the use of instrumentation, expensive materials, and cold chain requirements. Through an iterative process, we optimized viral lysis and RNA binding to and elution from glass fiber membranes. Protocol changes were evaluated by testing eluates in virus-specific real-time RT-PCRs (rRT-PCRs). Efficient, non-toxic viral lysis was achieved with a sucrose buffer including KCl, proteinase K and carrier RNA. Glass fiber membranes demonstrated concentration-dependent RNA binding of three arthropod-borne RNA viruses (arboviruses): dengue, chikungunya and Oropouche. Membrane binding was significantly increased in an acidic arginine binding buffer. For the clinical evaluation, 36 dengue virus (DENV)-positive serum samples were extracted in duplicate in the optimized protocol and results were compared to a commercial method. DENV RNA was successfully extracted from 71/72 replicates (98.6%) in the extraction packets, and rRT-PCR Ct values correlated between the techniques. Five clinical samples were selected to evaluate ambient-temperature storage up to 7 days on dried glass fiber membranes. DENV RNA was stable at 1, 3 and 7 days post extraction, with a mean difference in eluate RNA concentration of 0.14 log_10_ copies/μL. At a cost of $0.08 /sample, RNA extraction and storage packets address key limitations to available protocols and may increase capacity for molecular detection of RNA viruses.

---

RNA viruses are the largest group of human viral pathogens and the most common cause of emerging infectious disease outbreaks.^1, 2^ Clinical management and disease containment rely on accurate laboratory diagnosis. For many RNA viruses, molecular methods provide the most sensitive and specific acute-phase diagnostics^3-7^, and RNA extraction remains a crucial step in sample preparation that ensures optimal performance of such methods. However, extraction presents many challenges owing to the relative instability of RNA compared to DNA and the presence of RNA degrading enzymes (RNases) and PCR inhibitors in clinical samples.^8-10^ Extraction is generally performed using commercial kits that are costly and rely on proprietary materials that can be difficult to obtain in low-resource settings or emerging markets.^11, 12^ Kits often require the use of dedicated instruments, corrosive and hazardous chemicals, and -80°C storage of the resulting eluate if testing will not be performed within 24 hours.^8-10, 13, 14^ As a result, RNA extraction and storage remain major barriers to the implementation and use of molecular methods in low-resource settings.

Arboviruses comprise the subset of RNA viruses transmitted by infected arthropod vectors such as mosquitoes and ticks. These have resulted in large, recent outbreaks caused by the introduction of viruses into naïve populations [e.g., chikungunya virus (CHIKV) and Zika virus (ZIKV) in the Americas in 2014-2016]^15-18^ or re-emergence of viruses in populations residing in endemic regions [e.g., yellow fever virus (YFV) and dengue virus (DENV)].^19-23^ Of the arboviruses, DENV is responsible for the greatest burden of human disease, causing an estimated 100 million symptomatic infections (dengue cases) per year spread over 125 countries.^24^ Dengue presents with nonspecific, systemic symptoms that cannot be clinically differentiated from other causes of an acute febrile illness, and diagnostic confirmation relies on the availability of laboratory tests.^7, 20^ However, in endemic countries such as Paraguay, DENV causes large seasonal outbreaks that exhaust laboratory reagent supply and testing capacity, resulting in under detection and potentially worse clinical outcomes.^4, 25, 26^

In this study, we sought to address barriers to RNA virus detection that result from available extraction methods and the inherent challenges of working with RNA. We developed a simple, safe, and economical protocol for RNA extraction and storage from serum and plasma in resource-limited settings. The protocol was developed and optimized using contrived DENV clinical samples and RNA from species representing the 3 predominant genera of arboviruses: DENV, a flavivirus; CHIKV, an alphavirus; and Oropouche virus (OROV), an orthobunyavirus. Clinical evaluation of the optimized protocol was then performed on a set of 36 acute-phase samples from confirmed dengue cases in Paraguay.

## Results

### Viral lysis

Four experimental lysis buffers (deionized water, STET (8% Sucrose, 5% Triton^™^ X-100, 50mM Tris-HCl, and 50 mM EDTA), Sodium Dodecyl Sulfate-NaCl, and sucrose buffer) were evaluated for the ability to lyse DENV, followed by completion of RNA extraction in a commercial spin column protocol (QIAamp Viral RNA Mini Kit, Qiagen). SDS-NaCl and sucrose solutions performed similarly, yielding earlier DENV cycle threshold (Ct) values (indicating increased RNA yield) by rRT-PCR (Table S1). To eliminate potential SDS inhibition of downstream molecular testing, sucrose buffer was chosen as the lysis buffer for further experiments. To further enhance RNA recovery and prevent degradation, varying amounts of poly-A carrier RNA and proteinase K were added to the lysis buffer. In side-by-side comparisons, carrier RNA (2.5μg/sample) and proteinase K (5.0μg/sample) independently increased RNA recovery. Higher concentrations of carrier RNA and proteinase K in the lysis mixture did not enhance RNA recovery (5 and 10μg/sample, respectively; Tables S2 and S3). In addition, the impact of lysis incubation on RNA recovery at room temperature was tested at various time durations from 1 to 60 minutes. Samples were stable for up to one hour in lysis mixture, however, longer incubation times did not result in increased RNA recovery after ten-minute sample incubation at room temperature (data not shown). Based on these data, a ten-minute incubation period was selected for the final procedure.

### Membranes

Extraction packets were assembled as shown in Figure 1. Whatman 3, Fusion 5, and glass fiber (GF/D) membranes were evaluated as the RNA binding membrane. To compare RNA recovery from the different membranes, 15μL of purified DENV, CHIKV, and OROV RNA were mixed with lysis buffer and ethanol and added to packets containing one of the 3 membranes. RNA was then eluted and tested by rRT-PCR. RNA recovery was successful for all viruses and concentrations on the GF/D membranes (12/12), whereas one extraction failed with both the Whatman 3 and Fusion 5 membranes (11/12 each; Table 1). Ct values were also lowest for RNA recovered from GF/D membranes, and based on these data, the GF/D membrane was selected for inclusion in the final RNA extraction packet.

**Table 1.**
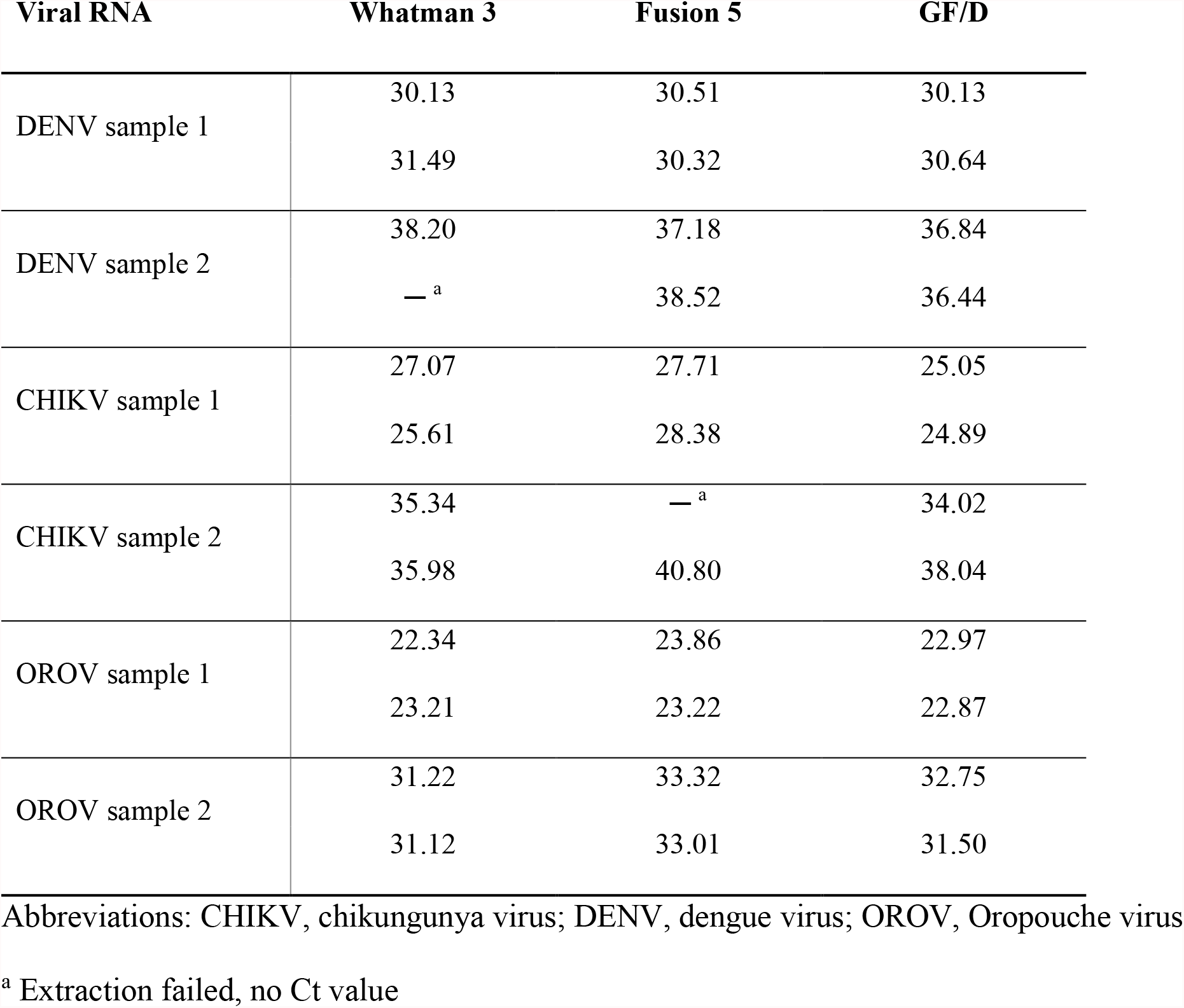
rRT-PCR Ct values for RNA of different arboviruses following binding to and elution from Whatman 3, Fusion 5, and GF/D membranes.

| **Viral RNA** | **Whatman 3** | **Fusion 5** | **GF/D** |
| --- | --- | --- | --- |
| DENV sample 1 | 30.13 | 30.51 | 30.13 |
|  | 31.49 | 30.32 | 30.64 |
| DENV sample 2 | 38.20 | 37.18 | 36.84 |
|  | —<sup>a</sup> | 38.52 | 36.44 |
| CHIKV sample 1 | 27.07 | 27.71 | 25.05 |
|  | 25.61 | 28.38 | 24.89 |
| CHIKV sample 2 | 35.34 | —<sup>a</sup> | 34.02 |
|  | 35.98 | 40.80 | 38.04 |
| OROV sample 1 | 22.34 | 23.86 | 22.97 |
|  | 23.21 | 23.22 | 22.87 |
| OROV sample 2 | 31.22 | 33.32 | 32.75 |
|  | 31.12 | 33.01 | 31.50 |

Abbreviations: CHIKV, chikungunya virus; DENV, dengue virus; OROV, Oropouche virus
<sup>a</sup> Extraction failed, no Ct value

**Figure 1.**
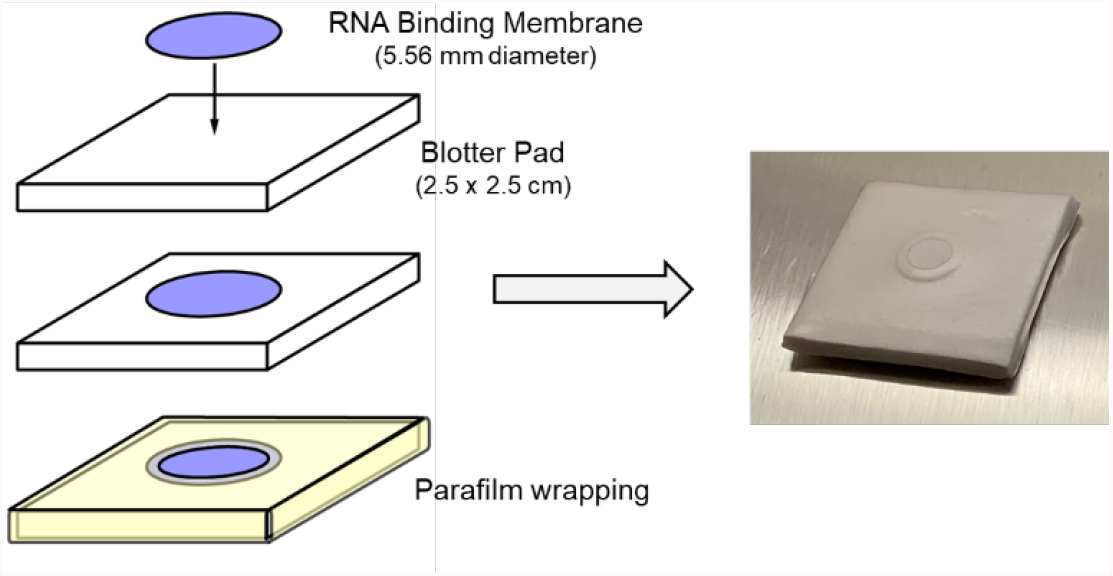
Components and assembly of RNA extraction packets (left) and an assembled packet (right). Whatman 3, Fusion 5, and GF/D were evaluated as RNA binding membranes.

### Amino acid binding buffers

Arginine- and glutamine-based amino acid binding buffers were assessed as a method to modulate RNA-membrane interactions when the lysate was loaded on the packet. Contrived DENV-positive serum samples were lysed and then treated with either amino acid buffer. Although results did not differ significantly, arginine treatment yielded increased RNA recovery, with lower Ct values overall and decreased variability (Figure S1). Based on these data, arginine was chosen for future experiments. To further evaluate the impact of arginine binding buffer, DENV, CHIKV and OROV RNA were added to the membranes in either the sucrose lysis buffer or an arginine buffer (plus ethanol in both cases). Relative to sucrose buffer, the arginine buffer demonstrated lower mean Ct values and decreased variability in Ct values with each membrane (Figure 2A). Ct values were significantly lower with the use of arginine buffer when data from all membranes were evaluated together (Figure 2A), and the relative increase in RNA yield is shown in Figure 2B.

**Figure 2.**
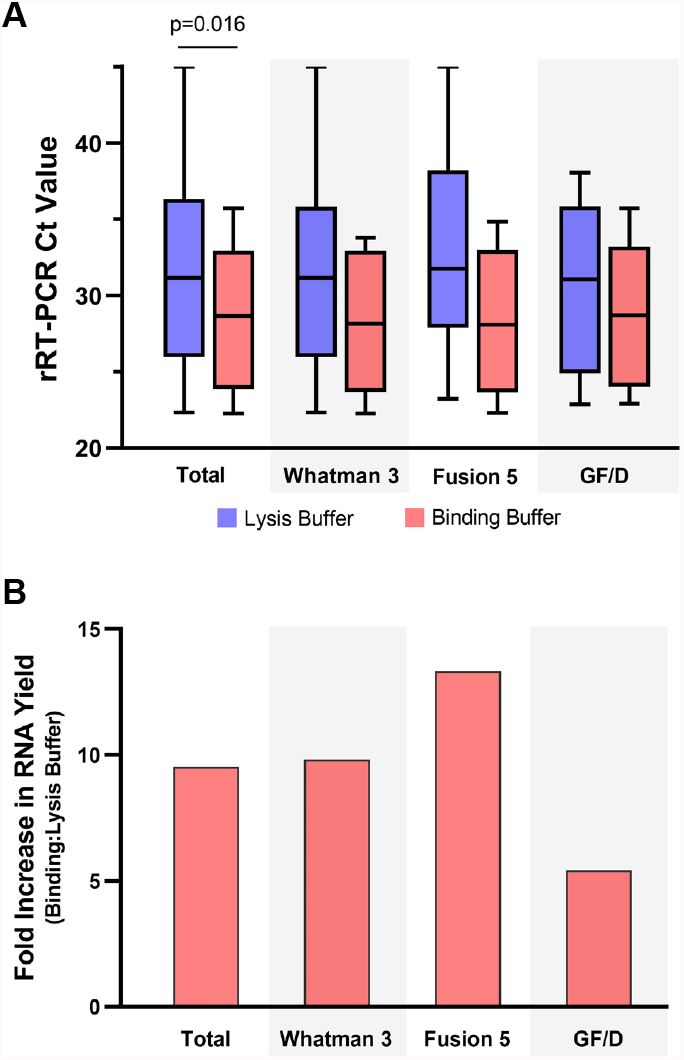
Amino acid binding buffers improve RNA yield from RNA extraction packets. **A**) Arginine binding buffer resulted in lower and more consistent rRT-PCR Ct values (improved RNA yield) following DENV, CHIKV and OROV RNA binding to and elution from extraction packets assembled with Whatman 3, Fusion 5, and GF/D membranes. Ct values were significantly lower when results from all membranes were pooled (Total; unpaired t-test); comparisons for individual membranes did not reach statistical significance. Box-and-whisker plots display median and range of all values. **B**) Fold increase in RNA yield with the use of arginine binding buffer compared to lysis buffer. Estimated based on expected 3.3 cycle *decrease* in Ct value for each 10-fold *increase* in RNA concentration in an rRT-PCR with 100% efficiency.

Following the initial analytical evaluation with purified RNA, arginine buffer was evaluated as a viral lysis buffer in place of sucrose buffer. However, testing with contrived DENV samples resulted in worse RNA recovery (Figure S2A). The arginine buffer was subsequently integrated as a binding buffer that is added after incubation of the sample in lysis buffer. This allowed for both successful viral lysis and improved binding of viral RNA to the packet membrane (Figure S2B). With the addition of the arginine buffer to the procedural workflow, MgCl_2_ in the sucrose buffer was changed to KCl to harmonize buffer preparations, which had no impact on RNA recovery (Table S4).

### Final workflow

Figure 3 shows the final extraction packet workflow. 25μL of serum or plasma is added to a 1.5mL tube with 25μL lysis mixture (2.5μL carrier RNA, 5μL proteinase k and 17.5μL lysis buffer). Following ten-minute incubation at ambient temperature, 100μL of amino acid binding buffer and 150μL of 90% ethanol are added to the lysate and mixed thoroughly by pipette. The resulting mixture is loaded dropwise onto the packets. Membranes are subsequently washed with 100μL glycine buffer and transferred by pipette tip to a tube containing 50μL of TE elution buffer. Membranes are incubated in elution buffer for 1 min and then discarded.

**Figure 3.**
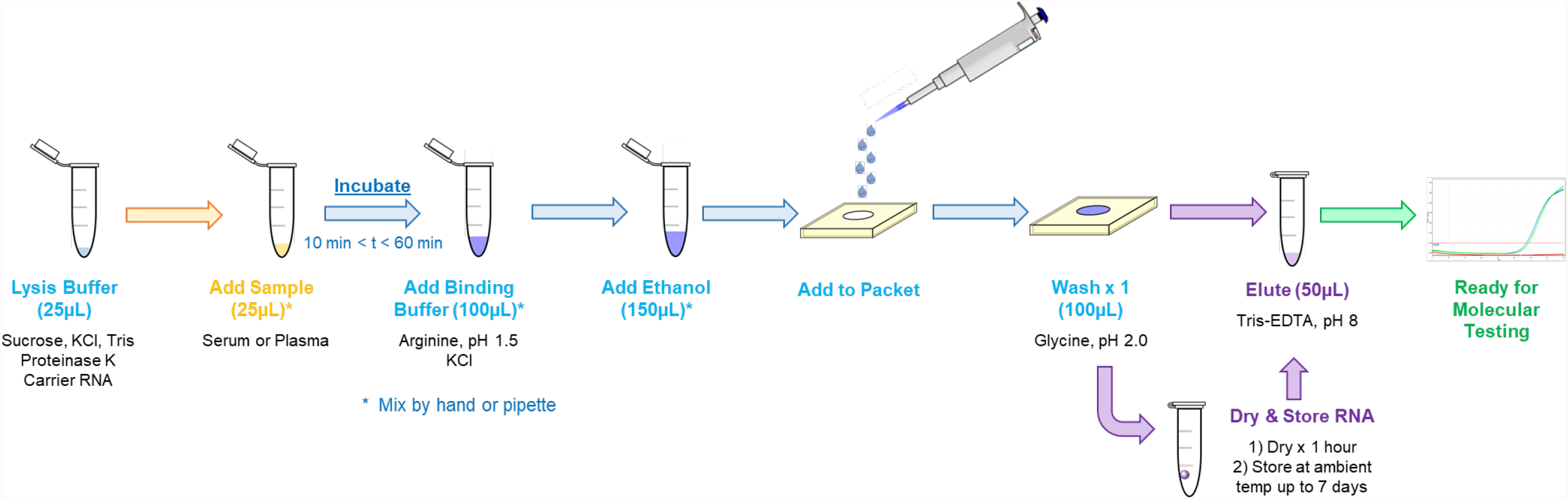
Optimized packet workflow for RNA extraction and storage. All steps are performed at ambient temperature.

### Clinical evaluation

36 DENV-positive serum samples from Paraguay, collected from 2018-2020, were selected for the clinical evaluation of the extraction packets. Demographic information and laboratory data are shown in Table 2. Samples included serotypes DENV-1 (20/36, 55.6%) and DENV-4 (16/36, 44.4%), which were the predominant DENV serotypes in Paraguay during those years. Viral loads ranged from 4.73 to 8.22 log_10_ copies/mL of serum. Samples were extracted in duplicate with the extraction packets (72 total extractions) and once in an EMAG robotic extraction instrument. DENV RNA was successfully extracted from 71/72 replicates (98.6%) in the extraction packets, and DENV multiplex rRT-PCR Ct values correlated with results following EMAG extraction (Figure 4, Table S5).

**Table 2.** Clinical and DENV laboratory data for 36 clinical samples extracted with the RNA packets.

| Category | Result |
| --- | --- |
| Total, n | 36 |
| Gender, female, n (%) | 21 (58.3) |
| Age, mean (SD) | 28.9 (14.1) |
| Day post-symptom onset, mean (SD) | 3.6 (1.6) |
| Serotype, n (%) |  |
| DENV-1 | 20 (55.6) |
| DENV-4 | 16 (44.4) |
| Viral load, mean (SD) <sup>a</sup> | 7.1 (1.4) |

Abbreviations: DENV, dengue virus; SD, standard deviation
<sup>a</sup> Viral load expressed as log<sub>10</sub> copies/mL serum

**Figure 4.**
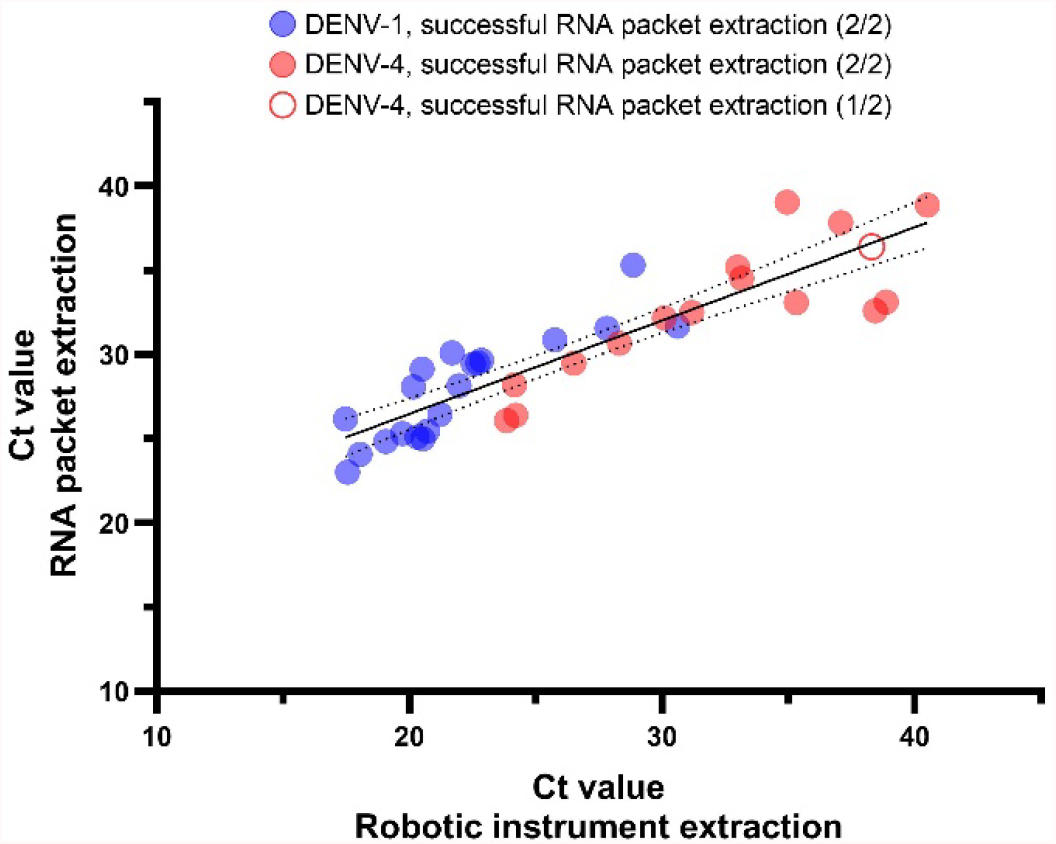
DENV RNA was successfully extracted from clinical samples using economical packets and rRT-PCR Ct values correlated with results following EMAG robotic extractions. Successful RNA extraction was defined as a positive result in the DENV multiplex rRT-PCR.^42, 43^ Average Ct values from duplicate packet extractions are graphed versus the Ct following EMAG extraction for 36 serum samples positive for DENV-1 (n=20) and DENV-4 (n=16). 71/72 eluates (98.6%) from the packets had detectable DENV RNA. The sample from which 1 of 2 replicates was positive is displayed as a half-filled circle.

Five DENV-1 samples were then selected for evaluation of RNA stability when stored on GF/D membranes at room temperature for up to 7 days. RNA was eluted on days 1, 3 and 7, and DENV RNA concentration in the eluates was calculated in the DENV multiplex rRT-PCR. Concentrations (Figure 5A) and Ct values (Figure S3) are shown for each time point. The median DENV RNA concentration in the eluates was 4.43 log_10_ copies/μL on day 0 and 4.57 log_10_ copies/μL on day 7 (0.14 log_10_ copies RNA/μL difference). DENV RNA concentration fluctuated between time points but remained within the expected error for quantitative rRT-PCR (±0.5 log_10_ copies, Figure 5B).

**Figure 5.**
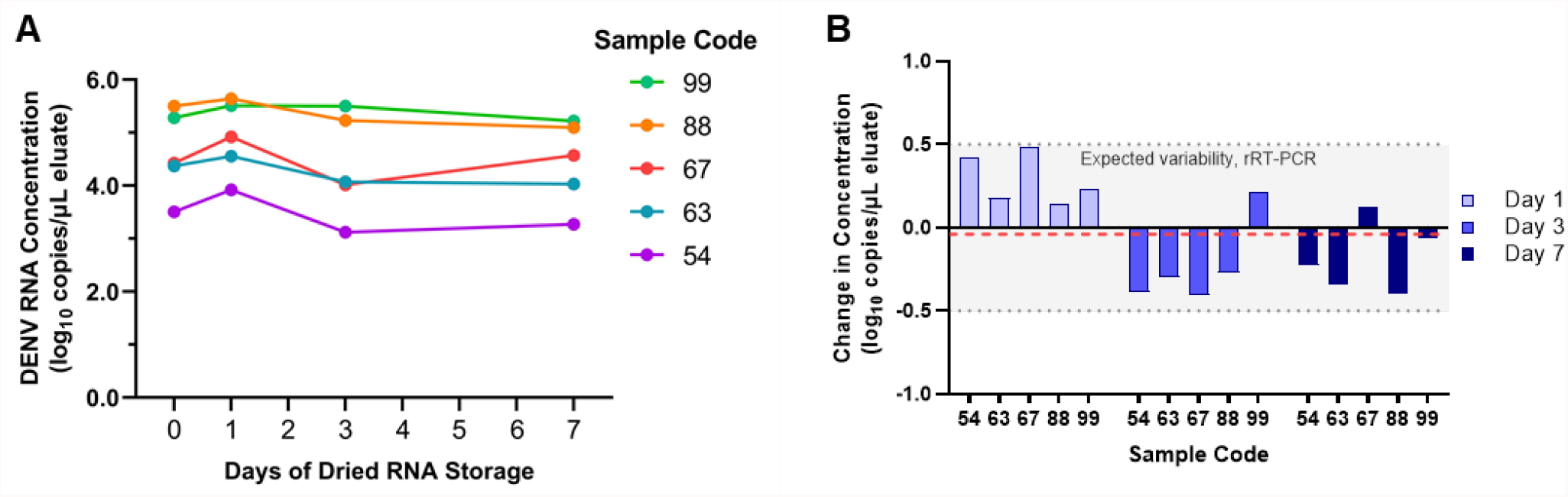
DENV RNA stored on dried extraction membranes is stable at ambient temperature for up to 7 days. **A**) Average DENV-1 RNA concentration in eluates from RNA extraction packets performed in duplicate is displayed for 5 clinical samples that were fully extracted on day 0 or that underwent lysis, addition to GF/D membrane-containing packets, glycine wash, drying and storage at ambient temperature. RNA was then eluted off the dried membranes and tested by rRT-PCR on days 1, 3 and 7. DENV RNA concentration was calculated from a 4-point standard curve included on each run. **B**) Change in RNA concentration between day 0 and days 1, 3, and 7. The shaded region highlights the expected intra-run variability of rRT-PCR (±0.5 log_10_ copies/μL). Mean change in concentration across time points (−0.04 log_10_ copies/μL) is displayed (red dashed line).

## Discussion

This study presents the development of an alternative, low-cost extraction method compatible with downstream rRT-PCR analysis for arboviral detection. Using the basic packet design of a previous method for viral DNA detection^27^, we developed and optimized a protocol that greatly improved extraction efficiency for RNA viruses. Our protocol is suitable for implementation in resource-limited settings as it eliminates the need for expensive proprietary materials, hazardous chemicals, or electricity during the extraction process. The final extraction packet protocol has an estimated cost of $0.08 /sample and successfully detected 98.7% of DENV-positive clinical samples when compared to a commercial robotic extraction system that has an initial cost of USD $115,000. Clinical samples were collected during two large DENV outbreaks in Paraguay (DENV-1 in 2018, DENV-4 in 2019-2020). Notably, the range of viral loads in samples that were available for extraction in the current study represent 92.7% of all DENV-positive samples with quantifiable viral loads from these two large outbreaks (Ref. 28, DENV-1; manuscript in progress, DENV-4). Samples with viral loads outside of this range were no longer available for extraction. These data indicate the potential utility of extraction packets in a real-world testing scenario outside of a high resource research laboratory.

In addition to examining the limitations posed by RNA extraction methods, in this study we address the temperature constraints of common RNA storage procedures. Existing methodologies require ultra-low storage temperatures that are costly to maintain and have limited capacity.^8, 13, 14^ In addition, the need for cold chain storage complicates the shipment of samples from collection sites to reference laboratories. We report an alternative ambient temperature technique where viral RNA is stored directly on the extraction membrane and integrity is maintained for downstream rRT-PCR analysis. This technique demonstrated RNA stability for up to 7 days post viral lysis for 5/5 clinical samples. Success of this technique likely results from washing with highly acidic buffers and drying the membrane, which serve to reduce RNA transesterification and RNase activity.^28, 29^ The developed packets, therefore, address major concerns for specimen preparation and provide a robust alternative for extraction and short-term storage of RNA.

To-date, there are few RNA extraction methods suitable for implementation in low-resource communities, none of which evaluate a system comparable to the extraction packet design.^30-32^ Various efforts to provide a sustainable alternative to expensive commercial kits utilize magnetic bead technology or solid-phase extraction methods that include biohazardous reagents. Magnetic bead technologies provide adjustable surface chemistries and ease of use for nucleic acid isolation.^32^ However, beads are relatively expensive and may be difficult to acquire and implement in limited resource settings. Other solid phase alternatives rely on toxic phenol and/or guanidine-based solutions that inhibit downstream molecular testing and require special handling in the laboratory.^33-35^ To both reduce inhibitory reagents in the procedure and utilize safer reagents for distribution outside of standard molecular laboratories, we assessed an alternative sucrose-based lysis buffer. Sucrose solutions have been used in various DNA and RNA isolation protocols and have demonstrated a key role in increasing yield of extracted nucleic material and RNA stability, without compromising testing integrity.^36, 37^ The RNA packet extraction protocol provides a simple alternative that relies on easily accessible reagents, including proteinase K, and can be prepared on site in a field setting.

In addition to the sucrose buffer, amino acid buffers were also examined as alternative lysis solutions to be used in lieu of chaotropic salts. A previous study found that several amino acid solutions provide favorable alternatives to standard buffers for nucleic acid recovery due to interactions between the amino acid, RNA and silica surface.^38^ Positively charged arginine and polar uncharged glutamine were chosen for evaluation within the extraction packet protocol for their distinct chemical properties. In the RNA extraction packets, low-pH arginine buffer improved RNA yield. However, promising analytical findings did not translate to improved DENV RNA recovery from contrived samples when arginine buffer replaced the optimized sucrose buffer in our extraction protocol. A combined protocol, integrating the arginine solution as a binding buffer after lysis in sucrose buffer, leveraged the properties of both solutions. These data demonstrate the applicability of amino acid buffers in RNA extraction protocols and highlight the importance of rigorous clinical evaluation for findings obtained under optimal laboratory conditions.

Limitations of the current study include the use of serum and plasma with the extraction packets. These typically require centrifugation for preparation, though represent the most common specimen types for DENV diagnostic testing.^4, 7, 13, 39, 40^ The use of carrier RNA and proteinase K within the lysis mixture also require cold storage following reconstitution, but both reagents can be shipped and stored in a lyophilized format. To address these limitations, future studies should evaluate adaptations to the extraction packets for use with whole blood and alternative methods for ambient temperature storage of carrier RNA and proteinase K.

Access to reliable and economical RNA extraction methods remains a significant barrier to viral molecular detection and surveillance. The packets developed during this study provide safe, economical, and reproducible RNA extraction from clinical samples. These RNA extraction and storage packets address key limitations to available protocols and may increase capacity for molecular detection of RNA viruses.

## Methods

### Packet design

The initial physical design of the packet was based on the filtration isolation of nucleic acids (FINA) method, as previously described.^27^ Basic packet design consisted of a 5.56mm diameter membrane disk, sandwiched between a square blotter pad base (2.5 × 2.5 cm; VWR International, Radnor, PA) and a Parafilm cover with a 3.96mm diameter opening (Research Products International, Mt. Prospect, IL) centered directly above the membrane disk (Figure 1). Packets were assembled with Whatman 3, Fusion 5, and glass microfiber (GF/D) membranes (all from MiliporeSigma, Burlington, MA). The initial extraction protocol consisted of 1) incubating 25μL of serum/plasma with a lysis mixture for 10 minutes, 2) addition of ethanol, 3) addition of the lysate-ethanol mixture to the extraction packet, 4) a single wash with 100μL 10X glycine solution (pH 2.7; Polysciences, Warrington, PA), and 5) elution of RNA by transfer of the membrane to a 1.5mL tube containing 50μL10mM TE buffer, pH 8.0 (Teknova, Hollister, CA). Membranes were incubated in TE for one minute and subsequently removed and disposed. Eluates were tested immediately by rRT-PCR or stored at 4°C for up to 24 hours.

### Lysis and binding buffers

Four experimental lysis buffers were evaluated: deionized water, STET (8% Sucrose, 5% Triton^™^ X-100, 50mM Tris-HCl, and 50 mM EDTA; Teknova, Hollister, CA), Sodium Dodecyl Sulfate-NaCl,^34^ and a sucrose buffer.^36^ Lysis buffers were initially incorporated into and evaluated with a membrane-based commercial protocol (QiaAMP Viral RNA Mini Kit, Qiagen, Germantown, MD). Contrived DENV samples were either lysed with buffer AVL (as part of the kit) or an experimental lysis buffer and then extracted with the remaining steps in the manufacturer’s protocol. Based on these experiments, the sucrose solution was chosen for the lysis buffer. Poly-A carrier RNA (2.5μg/sample, Qiagen, Germantown, MD) and proteinase K (5μg/sample; New England Biolabs, Ipswich, MA) were evaluated as additional components to the lysis mixture. Further experiments used 25μL/reaction of lysis mixture containing 17.5μL of lysis buffer, 2.5μL (2.5μg) carrier RNA and 5μL (5.0μg) proteinase K. All sucrose solution components were adjusted in an iterative manner for further optimization. NaCl, MgCl_2_, and KCl were evaluated as different chaotropic salts across a range of concentrations (50mM to 400mM) alongside varying sucrose concentrations (50mM to 300mM). Solution pH was also evaluated at neutral and slightly alkaline pH values (7.0, 7.5, and 8.0). The optimized sucrose lysis buffer contained 100mM sucrose (1.2 M; Boston BioProducts, Ashland, MA); 50mM Tris-HCL, pH 7.0; 100mM KCl (both from MilliporeSigma, USA).

Arginine and glutamine amino acid binding buffers were initially prepared as described.^41^ The buffer contained 100mM L-arginine (MilliporeSigma, USA) and 400 mM KCl. Binding buffer pH was optimized across a range from 1.5-9.1, with a final buffer pH of 1.5. Lysis and binding buffers were stored at room temperature. Following preparation, the lysis mixture was used immediately or stored at 4°C until use.

### Clinical samples and RNA stability

DENV-positive clinical samples were collected as part of an ongoing study to detect and characterize arboviral infections in Asunción, Paraguay in collaboration with the Instituto de Investigaciones en Ciencas de la Salud, Universidad Nacional de Asunción (IICS-UNA).^4^ Samples were selected for the current study that had detectable and quantifiable DENV viral loads in the DENV multiplex rRT-PCR and had sufficient volume remaining for re-extraction (100μL). All samples had been shipped to Emory on dry ice, stored at -80°C, and thawed at 4°C immediately prior to extraction. For the clinical evaluation, side-by-side extractions were performed with the extraction packets in duplicate and on an EMAG robotic extraction instrument (bioMérieux, Durham, NC). RNA was extracted from 25μL of sample and eluted into 50μL for both the packets and comparator extraction protocols. Eluates were tested immediately by rRT-PCR.

For the stability study, 5 DENV-1 samples with sufficient remaining volume were individually re-extracted with the packets to evaluate RNA stability on days 0, 1, 3, and 7 post extraction. For this evaluation, 8 extraction packets were prepared, with duplicate packets for each time point. On day 0, DENV RNA was completely extracted, eluted off two membranes, and run in the DENV multiplex rRT-PCR to establish a baseline reference. For the remainder of the time points, DENV RNA was extracted with the packets through the glycine wash step and transferred to empty 1.5mL tubes to air dry at ambient temperature for 1 hour. After drying, tubes were closed and stored in airtight plastic bags with desiccant packets. RNA was eluted from dried membranes with 50μL TE buffer on days 1, 3, and 7 following extractions. Eluates were run in the DENV multiplex rRT-PCR for comparison with day 0 results. A four-point standard curve was included on each run to calculate DENV-1 RNA concentration at each time point.^42, 43^

### Reference viral RNAs and contrived samples

Packet membranes were initially evaluated with viral RNA from DENV, CHIKV and OROV. DENV RNA for this portion of the study was a 135-base synthesized DENV-2 RNA oligonucleotide containing the DENV multiplex rRT-PCR target sequence (Ultramer RNA, Integrated DNA Technologies, Coralville, IA). For CHIKV and OROV, previously extracted (EMAG) genomic RNA was used. Contrived clinical samples were prepared by spiking negative human serum or plasma (MilliporeSigma, USA) with DENV-positive serum of known concentrations. Aliquots of contrived specimens were prepared and stored at -80°C until use.

### rRT-PCR

Eluates from optimization and analytical evaluation experiments were tested in a single-reaction multiplex rRT-PCR for ZIKV, CHIKV, and DENV or a singleplex rRT-PCR for OROV, both performed as previously described.^44, 45^ For the clinical evaluation, eluates were tested in the DENV multiplex rRT-PCR, which is a serotype-specific assay for DENV detection and quantitation.^42, 43^ All rRT-PCR reactions were performed in 20μL reactions of the SuperScript III Platinum One-Step qRT-PCR kit (Thermo Fisher) containing 5μL of eluate and run on a Rotor-Gene Q instrument (Qiagen). Positive and negative controls were included on each run, and rRT-PCRs were analyzed and interpreted as previously described.^42-45^

### Statistics

Basic statistical analyses were performed in Excel (Microsoft, Redmond, WA). Ct values obtained with lysis buffer and arginine binding buffer were compared by unpaired student’s t test (GraphPad Prism version 9.2, GraphPad, San Diego, CA). Graphs were prepared with GraphPad and Excel.

## Data Availability

All data produced in the present study are available upon reasonable request to the authors.

## Acknowledgements

This research was supported by the Doris Duke Foundation (Clinical Scientist Development Award 2019089) and the National Institute of Allergy and Infectious Diseases (R21AI146443). We thank all members of the research team at the Instituto de Investigaciones en Ciencias de la Salud, Universidad Nacional de Asunción, and the participants and their family members who have contributed to ongoing studies of arboviral infections in Paraguay. We also thank Katherine Immergluck, Maxwell Su, and Victoria Stittleburg at Emory University for their assistance over the course of this project.

## Supplemental Material

**Figure S1.**
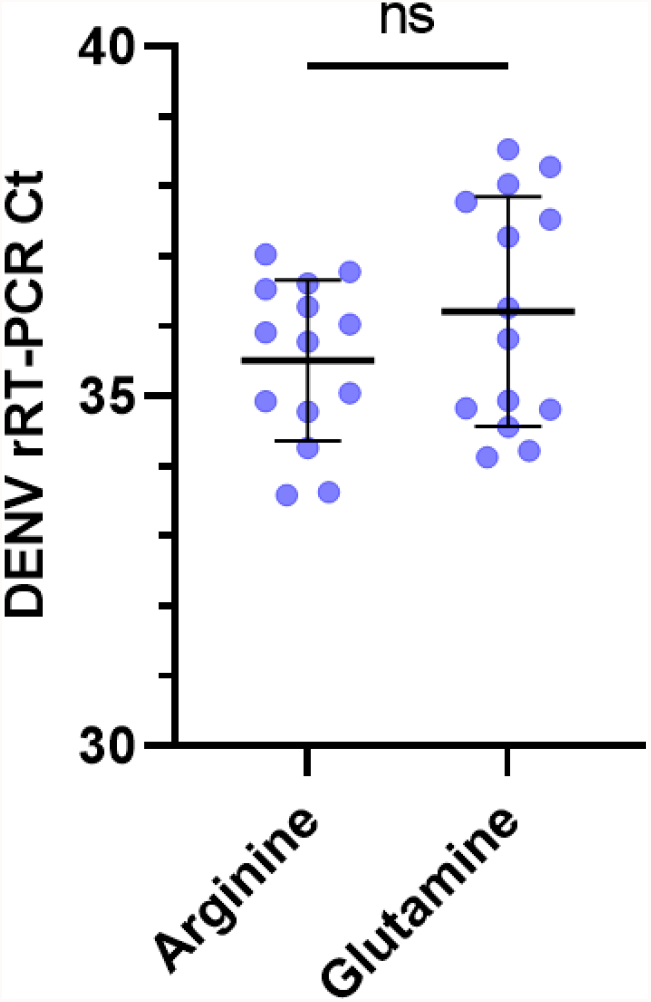
rRT-PCR cycle threshold (Ct) values for lysed DENV-positive serum samples treated with glutamine versus arginine binding buffer prior to addition to extraction packets. All evaluations were done with glass fiber GF/D membranes. Ct values were not significantly different for samples treated with arginine (mean, 35.51; standard deviation, 1.15) versus glutamine (36.21; 1.64; p = 0.2).

**Figure S2.**
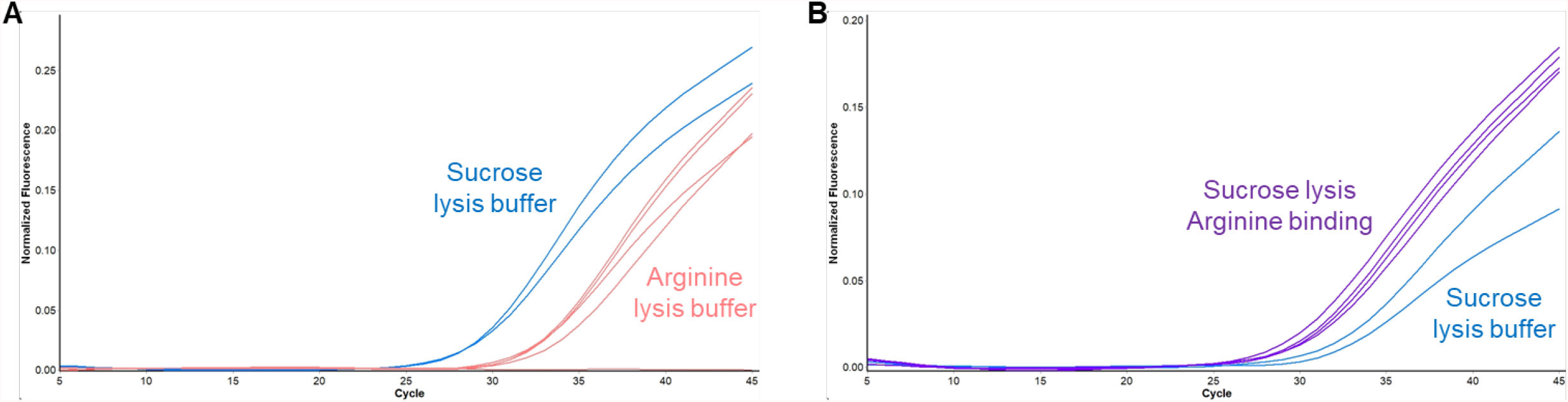
**A**) Use of arginine buffer for viral lysis results in reduced RNA yield from extraction packets. **B**) A combined protocol incorporating arginine buffer as a binding buffer after lysis in sucrose buffer results in increased RNA yield compared to an extraction protocol without a binding buffer. Different contrived samples were used on the two runs.

**Figure S3.**
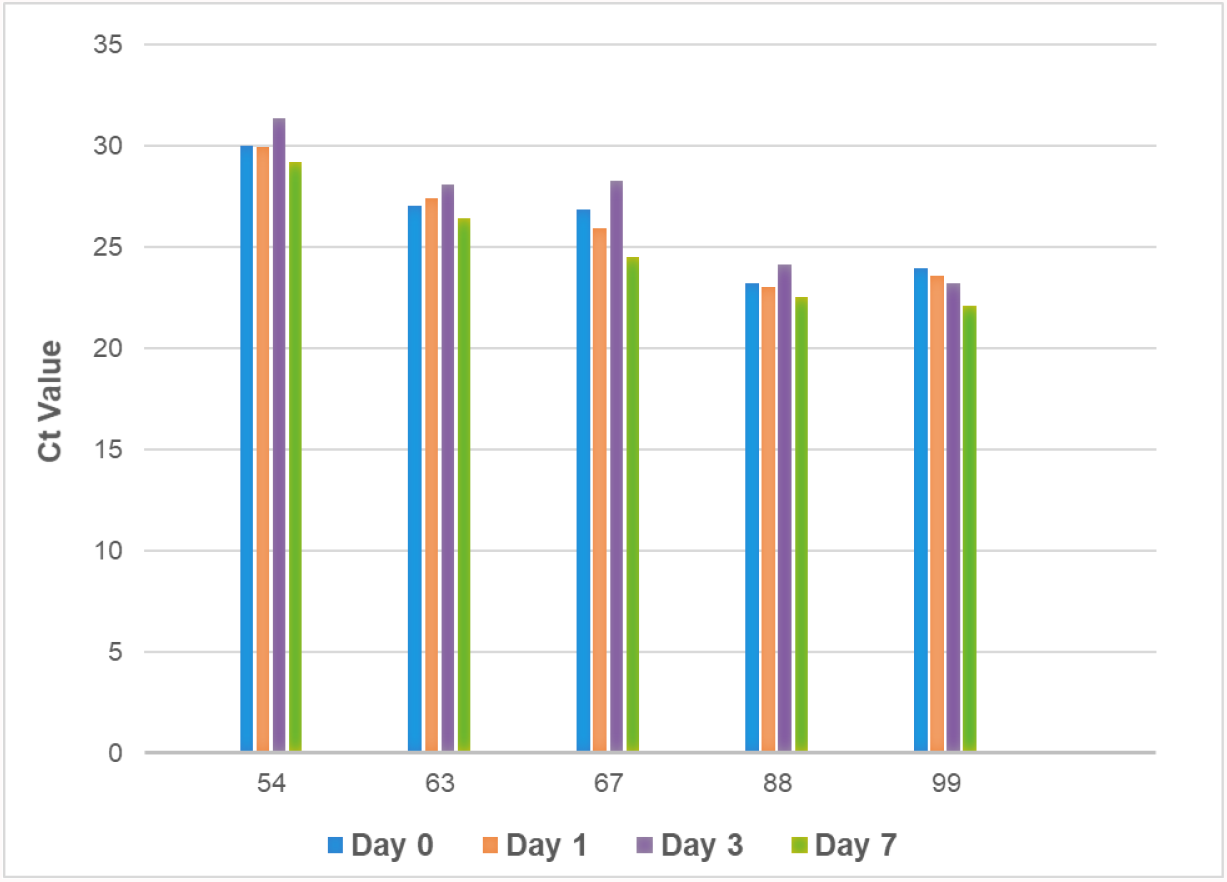
Stability of DENV Ct values from 5 clinical samples that were eluted from packets after complete extraction on day 0 or following storage at ambient temperature on the membranes prior to elution on days 1, 3, and 7. Ct values are averages of duplicate extractions tested in the DENV multiplex rRT-PCR. Sample numbers are displayed on the x-axis.

**Table S1.**
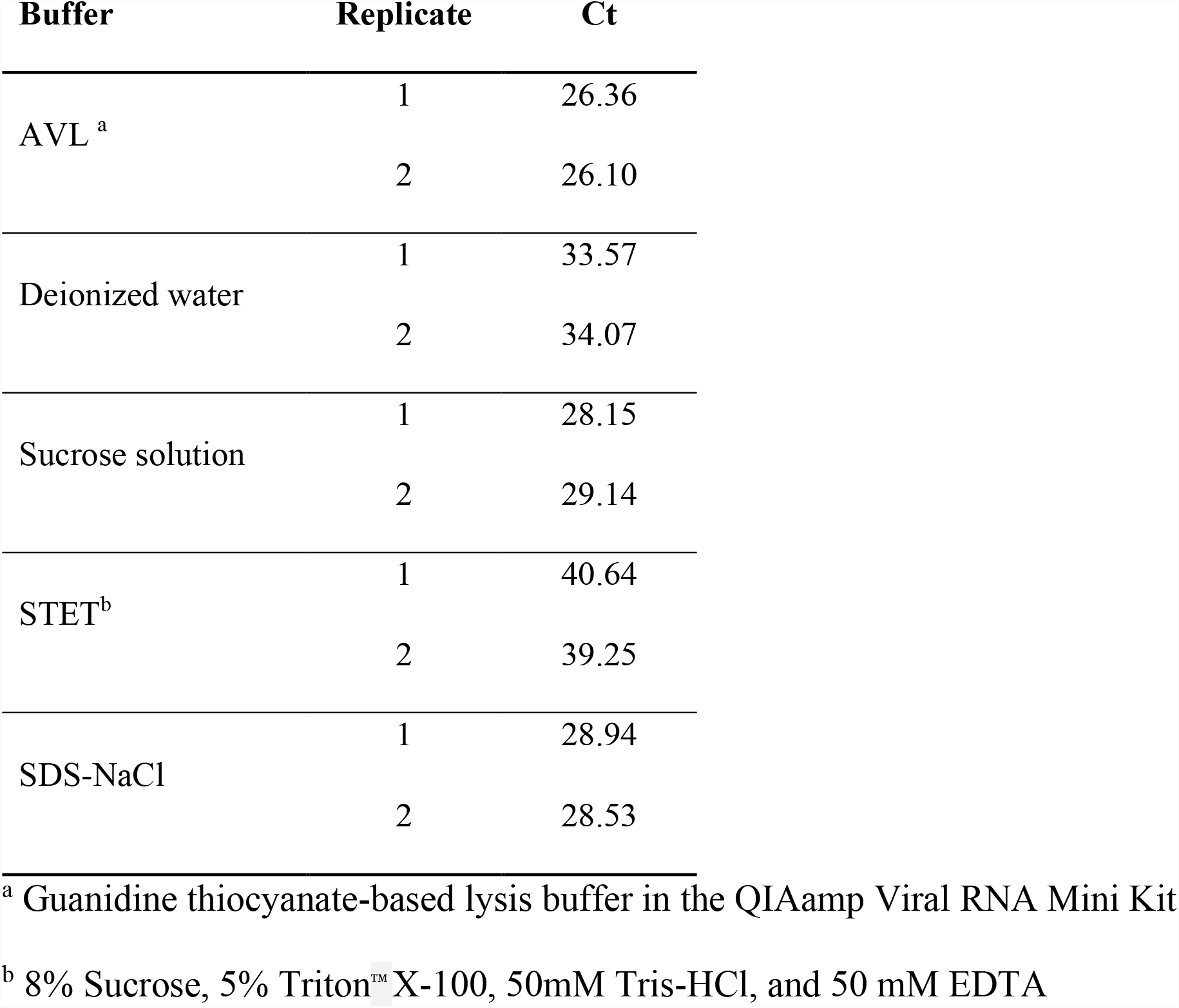
Comparison of DENV Ct values following RNA extraction in experimental lysis buffers.

| Buffer | Replicate | Ct |
| --- | --- | --- |
| AVL <sup>a</sup> | 1 | 26.36 |
|  | 2 | 26.10 |
| Deionized water | 1 | 33.57 |
|  | 2 | 34.07 |
| Sucrose solution | 1 | 28.15 |
|  | 2 | 29.14 |
| STET<sup>b</sup> | 1 | 40.64 |
|  | 2 | 39.25 |
| SDS-NaCl | 1 | 28.94 |
|  | 2 | 28.53 |

<sup>a</sup> Guanidine thiocyanate-based lysis buffer in the QIAamp Viral RNA Mini Kit
<sup>b</sup> 8% Sucrose, 5% Triton™ X-100, 50mM Tris-HCl, and 50 mM EDTA

**Table S2.** Comparison of DENV Ct values following RNA extraction in lysis mixtures with and without carrier RNA.

| **Replicate** | **Sucrose**<br>**buffer only** | **Carrier RNA**<br>**(2.5ug/sample)** | **Carrier RNA**<br>**(5.0ug/sample) <sup>a</sup>** |
| --- | --- | --- | --- |
| 1 | 29.30 | 27.27 | 27.10 |
| 2 | 30.13 | 26.75 | 27.79 |
| 3 | 30.27 | 27.27 | — |
| 4 | 30.08 | 26.75 | — |
| 5 | 30.51 | 27.03 | — |
| 6 | 29.72 | 27.42 | — |
| Average | 30.00 | 27.08 | 27.45 |

<sup>a</sup> “—” indicates not tested

**Table S3.** Comparison of DENV Ct values following RNA extraction in lysis mixtures with and without proteinase K.

| **Replicate** | **Sucrose buffer**<br>**+ carrier RNA** | **Proteinase K**<br>**(2.5ug/sample)** | **Proteinase K**<br>**(5.0ug/sample) <sup>a</sup>** |
| --- | --- | --- | --- |
| 1 | 32.09 | 28.34 | 28.38 |
| 2 | 29.50 | 27.12 | 27.24 |
| 3 | 31.14 | 27.31 | 27.38 |
| 4 | 33.00 | 26.23 | 26.92 |
| 5 | 32.46 | 26.48 | — |
| 6 | 32.71 | 26.35 | — |
| Average | 31.82 | 26.97 | 27.48 |

<sup>a</sup> “—” indicates not tested

**Table S4.** Comparison of DENV Ct values following extractions using sucrose lysis buffer containing 100mM MgCl_2_ or KCl.

| Sample | Replicate | <u>Sucrose Lysis Buffer</u> |  |
| --- | --- | --- | --- |
|  |  | MgCl<sub>2</sub> | KCl |
| 1 | 1 | 31.99 | 31.10 |
|  | 2 | 30.90 | 31.33 |
| 2 | 1 | 34.73 | 34.66 |
|  | 2 | 34.34 | 34.09 |

**Table S5.**
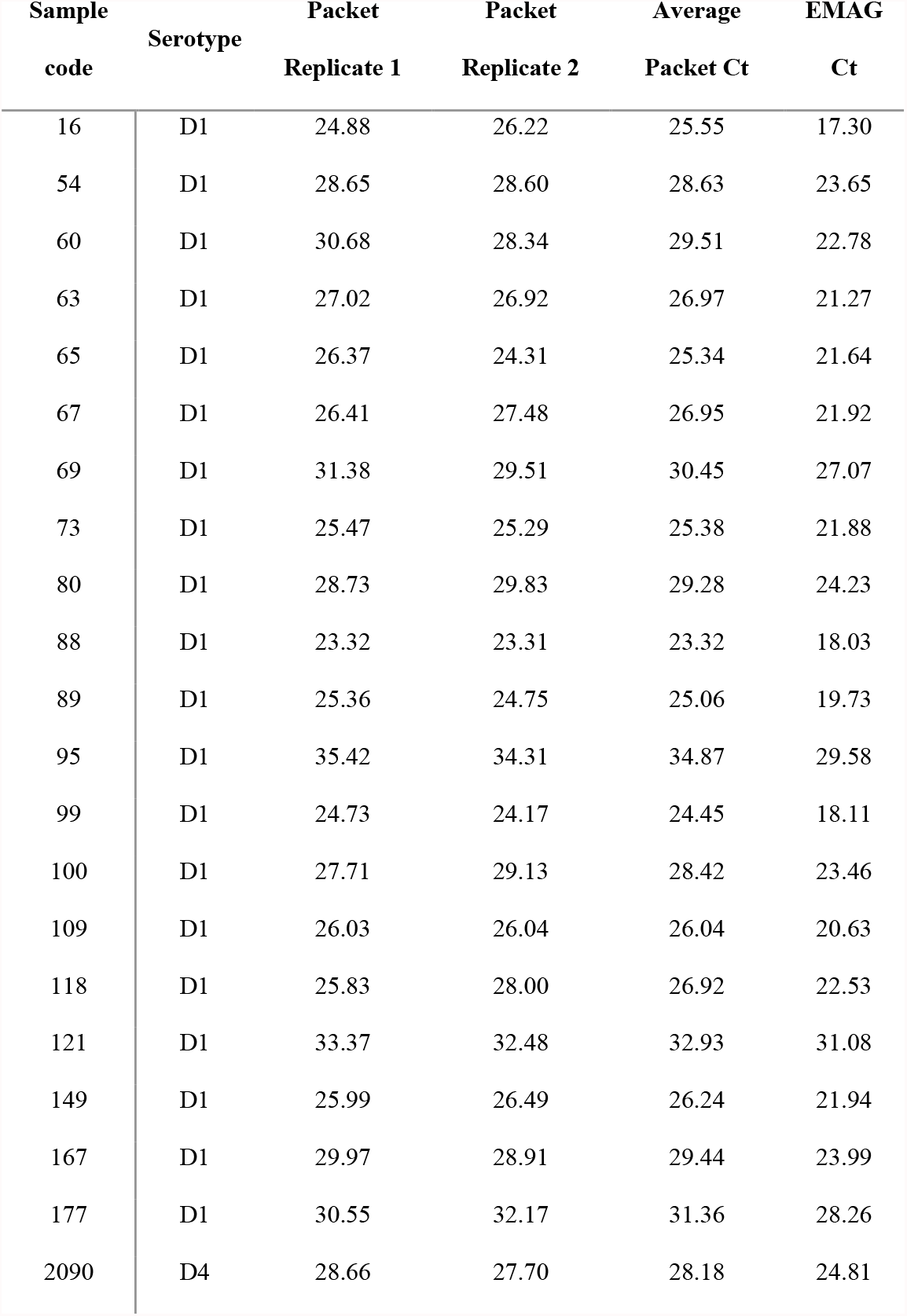
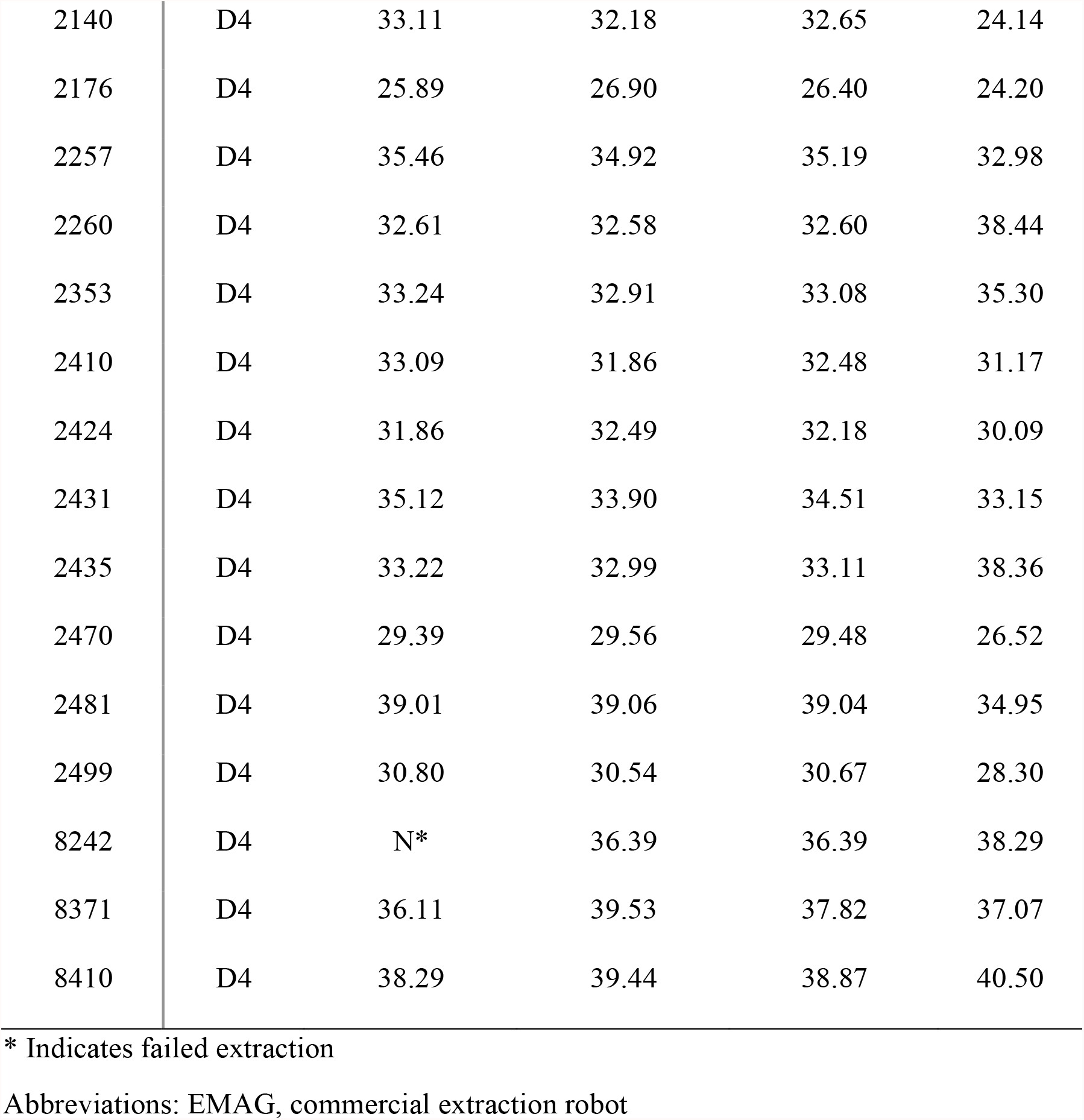
Ct values following RNA extraction with economical packets and a commercial robotic system.

| **Sample**<br>**code** | **Serotype** | **Packet**<br>**Replicate 1** | **Packet**<br>**Replicate 2** | **Average**<br>**Packet Ct** | **EMAG**<br>**Ct** |
| --- | --- | --- | --- | --- | --- |
| 16 | D1 | 24.88 | 26.22 | 25.55 | 17.30 |
| 54 | D1 | 28.65 | 28.60 | 28.63 | 23.65 |
| 60 | D1 | 30.68 | 28.34 | 29.51 | 22.78 |
| 63 | D1 | 27.02 | 26.92 | 26.97 | 21.27 |
| 65 | D1 | 26.37 | 24.31 | 25.34 | 21.64 |
| 67 | D1 | 26.41 | 27.48 | 26.95 | 21.92 |
| 69 | D1 | 31.38 | 29.51 | 30.45 | 27.07 |
| 73 | D1 | 25.47 | 25.29 | 25.38 | 21.88 |
| 80 | D1 | 28.73 | 29.83 | 29.28 | 24.23 |
| 88 | D1 | 23.32 | 23.31 | 23.32 | 18.03 |
| 89 | D1 | 25.36 | 24.75 | 25.06 | 19.73 |
| 95 | D1 | 35.42 | 34.31 | 34.87 | 29.58 |
| 99 | D1 | 24.73 | 24.17 | 24.45 | 18.11 |
| 100 | D1 | 27.71 | 29.13 | 28.42 | 23.46 |
| 109 | D1 | 26.03 | 26.04 | 26.04 | 20.63 |
| 118 | D1 | 25.83 | 28.00 | 26.92 | 22.53 |
| 121 | D1 | 33.37 | 32.48 | 32.93 | 31.08 |
| 149 | D1 | 25.99 | 26.49 | 26.24 | 21.94 |
| 167 | D1 | 29.97 | 28.91 | 29.44 | 23.99 |
| 177 | D1 | 30.55 | 32.17 | 31.36 | 28.26 |
| 2090 | D4 | 28.66 | 27.70 | 28.18 | 24.81 |
| 2140 | D4 | 33.11 | 32.18 | 32.65 | 24.14 |
| 2176 | D4 | 25.89 | 26.90 | 26.40 | 24.20 |
| 2257 | D4 | 35.46 | 34.92 | 35.19 | 32.98 |
| 2260 | D4 | 32.61 | 32.58 | 32.60 | 38.44 |
| 2353 | D4 | 33.24 | 32.91 | 33.08 | 35.30 |
| 2410 | D4 | 33.09 | 31.86 | 32.48 | 31.17 |
| 2424 | D4 | 31.86 | 32.49 | 32.18 | 30.09 |
| 2431 | D4 | 35.12 | 33.90 | 34.51 | 33.15 |
| 2435 | D4 | 33.22 | 32.99 | 33.11 | 38.36 |
| 2470 | D4 | 29.39 | 29.56 | 29.48 | 26.52 |
| 2481 | D4 | 39.01 | 39.06 | 39.04 | 34.95 |
| 2499 | D4 | 30.80 | 30.54 | 30.67 | 28.30 |
| 8242 | D4 | N* | 36.39 | 36.39 | 38.29 |
| 8371 | D4 | 36.11 | 39.53 | 37.82 | 37.07 |
| 8410 | D4 | 38.29 | 39.44 | 38.87 | 40.50 |

\* Indicates failed extraction
Abbreviations: EMAG, commercial extraction robot

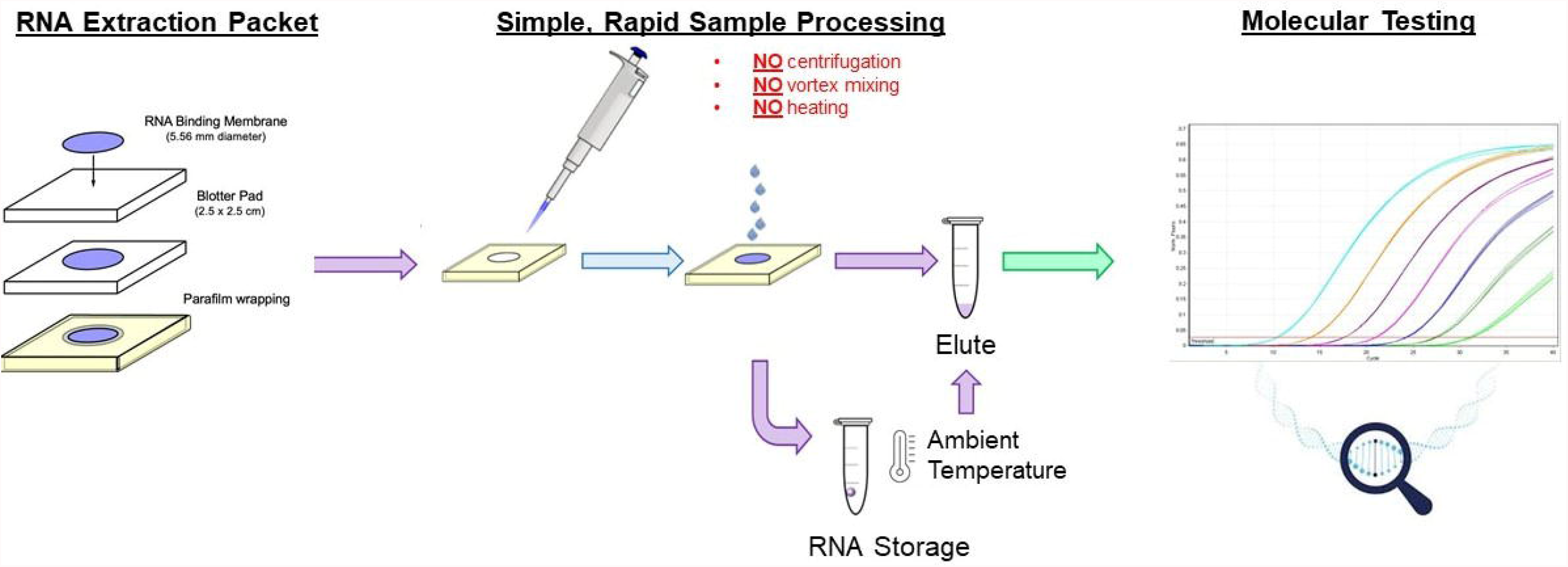

## References

1. Carrasco-Hernandez, R.; Jácome, R.; López Vidal, Y.; Ponce De León, S., Are RNA Viruses Candidate Agents for the Next Global Pandemic? A Review. ILAR Journal 2017, 58 (3), 343–358.

2. Rosenberg, R., Detecting the emergence of novel, zoonotic viruses pathogenic to humans. Cellular and Molecular Life Sciences 2015, 72 (6), 1115–1125.

3. Natrajan, M. S.; Rojas, A.; Waggoner, J. J., Beyond Fever and Pain: Diagnostic Methods for Chikungunya Virus. Journal of clinical microbiology 2019.

4. Rojas, A.; Cardozo, F.; Cantero, C.; Stittleburg, V.; Lopez, S.; Bernal, C.; Gimenez Acosta, F. E.; Mendoza, L.; Pinsky, B. A.; Arevalo de Guillen, I.; Paez, M.; Waggoner, J., Characterization of dengue cases among patients with an acute illness, Central Department, Paraguay. PeerJ 2019, 7, e7852.

5. Waggoner, J. J.; Pinsky, B. A., Zika Virus: Diagnostics for an Emerging Pandemic Threat. Journal of clinical microbiology 2016, 54 (4), 860–7.

6. Waggoner, J. J.; Rojas, A.; Pinsky, B. A., Yellow Fever Virus: Diagnostics for a Persistent Arboviral Threat. Journal of clinical microbiology 2018, 56 (10).

7. World Health Organization, Dengue: guidelines for diagnosis, treatment, prevention and control. WHO Press: France, 2009.

8. Fabre, A. L.; Colotte, M.; Luis, A.; Tuffet, S.; Bonnet, J., An efficient method for long-term room temperature storage of RNA. Eur J Hum Genet 2014, 22 (3), 379–85.

9. Paul, R.; Ostermann, E.; Wei, Q., Advances in point-of-care nucleic acid extraction technologies for rapid diagnosis of human and plant diseases. Biosens Bioelectron 2020, 169, 112592.

10. Tan, S. C.; Yiap, B. C., DNA, RNA, and protein extraction: the past and the present. J Biomed Biotechnol 2009, 2009, 574398.

11. Domingo, C.; Niedrig, M.; Teichmann, A.; Kaiser, M.; Rumer, L.; Jarman, R. G.; Donoso-Mantke, O., 2nd international external quality control assessment for the molecular diagnosis of Dengue infections. PLoS neglected tropical diseases 2010, 4 (10), e833.

12. Satyanarayana, M., Material shortages hamper testing for novel coronavirus. C&EN 2020, 98 (12), 6.

13. Centers for Disease Control and Prevention, CDC DENV-1-4 Real-Time RT-PCR Assay: Instructions for Use. 2013.

14. Centers for Disease Control and Prevention, Trioplex Real-time RT-PCR Assay: Instructions for Use. 2021.

15. Charrel, R. N.; Leparc-Goffart, I.; Gallian, P.; de Lamballerie, X., Globalization of Chikungunya: 10 years to invade the world. Clinical microbiology and infection : the official publication of the European Society of Clinical Microbiology and Infectious Diseases 2014, 20 (7), 662–3.

16. Weaver, S. C.; Lecuit, M., Chikungunya virus and the global spread of a mosquito-borne disease. The New England journal of medicine 2015, 372 (13), 1231–9.

17. Musso, D.; Gubler, D. J., Zika Virus. Clinical microbiology reviews 2016, 29 (3), 487–524.

18. Waggoner, J. J.; Pinsky, B. A., How great is the threat of chikungunya virus? Expert review of anti-infective therapy 2015, 13 (3), 291–3.

19. Bhatt, S.; Gething, P. W.; Brady, O. J.; Messina, J. P.; Farlow, A. W.; Moyes, C. L.; Drake, J. M.; Brownstein, J. S.; Hoen, A. G.; Sankoh, O.; Myers, M. F.; George, D. B.; Jaenisch, T.; Wint, G. R.; Simmons, C. P.; Scott, T. W.; Farrar, J. J.; Hay, S. I., The global distribution and burden of dengue. Nature 2013, 496 (7446), 504–7.

20. Guzman, M. G.; Harris, E., Dengue. Lancet 2015, 385 (9966), 453–65.

21. Shepard, D. S.; Undurraga, E. A.; Halasa, Y. A.; Stanaway, J. D., The global economic burden of dengue: a systematic analysis. The Lancet. Infectious diseases 2016.

22. Zhao, S.; Stone, L.; Gao, D.; He, D., Modelling the large-scale yellow fever outbreak in Luanda, Angola, and the impact of vaccination. PLoS neglected tropical diseases 2018, 12 (1), e0006158.

23. Dexheimer Paploski, I. A.; Souza, R. L.; Tauro, L. B.; Cardoso, C. W.; Mugabe, V. A.; Pereira Simoes Alves, A.B.; de Jesus Gomes, J.; Kikuti, M.; Campos, G. S.; Sardi, S.; Weaver, S. C.; Reis, M. G.; Kitron, U.; Ribeiro, G. S., Epizootic Outbreak of Yellow Fever Virus and Risk for Human Disease in Salvador, Brazil. Annals of internal medicine 2017.

24. Messina, J. P.; Brady, O. J.; Golding, N.; Kraemer, M. U. G.; Wint, G. R. W.; Ray, S. E.; Pigott, D. M.; Shearer, F. M.; Johnson, K.; Earl, L.; Marczak, L. B.; Shirude, S.; Davis Weaver, N.; Gilbert, M.; Velayudhan, R.; Jones, P.; Jaenisch, T.; Scott, T. W.; Reiner, R. C.; Hay, S. I., The current and future global distribution and population at risk of dengue. Nature Microbiology 2019, 4 (9), 1508–1515.

25. Rojas, A.; Aria, L.; de Guillen, Y. A.; Acosta, M. E.; Infanzón, B.; Diaz, V.; López, L.; Meza, T.; Riveros, O., Perfil clínico, hematológico y serológico en pacientes con sospecha de dengue del IICS-UNA, 2009-2013. Mem Inst Investig Cienc Salud 2016, 14 (2), 68–74.

26. Balmaseda, A.; Hammond, S. N.; Tellez, Y.; Imhoff, L.; Rodriguez, Y.; Saborio, S. I.; Mercado, J. C.; Perez, L.; Videa, E.; Almanza, E.; Kuan, G.; Reyes, M.; Saenz, L.; Amador, J. J.; Harris, E., High seroprevalence of antibodies against dengue virus in a prospective study of schoolchildren in Managua, Nicaragua. Tropical medicine & international health : TM & IH 2006, 11 (6), 935–42.

27. McFall, S. M.; Wagner, R. L.; Jangam, S. R.; Yamada, D. H.; Hardie, D.; Kelso, D. M., A simple and rapid DNA extraction method from whole blood for highly sensitive detection and quantitation of HIV-1 proviral DNA by real-time PCR. Journal of virological methods 2015, 214, 37–42.

28. Seyhan, A. A.; Burke, J. M., Mg2+-independent hairpin ribozyme catalysis in hydrated RNA films. RNA 2000, 6 (2), 189–98.

29. Perreault, D. M.; Ansly, E. V., Unifying the Current Data on the Mechanism of Cleavage -Transesterification of RNA. Angew. Clrem. Int. Ed. Eng. 1997, 36, 432–450.

30. Han, P.; Go, M. K.; Chow, J. Y.; Xue, B.; Lim, Y. P.; Crone, M. A.; Storch, M.; Freemont, P. S.; Yew, W. S., A high-throughput pipeline for scalable kit-free RNA extraction. Scientific Reports 2021, 11 (1).

31. Wozniak, A.; Cerda, A.; Ibarra-Henríquez, C.; Sebastian, V.; Armijo, G.; Lamig, L.; Miranda, C.; Lagos, M.; Solari, S.; Guzmán, A. M.; Quiroga, T.; Hitschfeld, S.; Riveras, E.; Ferrés, M.; Gutiérrez, R. A.; García, P., A simple RNA preparation method for SARS-CoV-2 detection by RT-qPCR. Scientific Reports 2020, 10 (1).

32. Pearlman, S. I.; Leelawong, M.; Richardson, K. A.; Adams, N. M.; Russ, P. K.; Pask, M. E.; Wolfe, A. E.; Wessely, C.; Haselton, F. R., Low-Resource Nucleic Acid Extraction Method Enabled by High-Gradient Magnetic Separation. ACS Applied Materials & Interfaces 2020, 12 (11), 12457–12467.

33. Yaffe, H.; Buxdorf, K.; Shapira, I.; Ein-Gedi, S.; Moyal-Ben Zvi, M.; Fridman, E.; Moshelion, M.; Levy, M., LogSpin: a simple, economical and fast method for RNA isolation from infected or healthy plants and other eukaryotic tissues. BMC research notes 2012, 5, 45.

34. Nwokeoji, A. O.; Kilby, P. M.; Portwood, D. E.; Dickman, M. J., RNASwift: A rapid, versatile RNA extraction method free from phenol and chloroform. Anal Biochem 2016, 512, 36–46.

35. Seok, Y.; Batule, B. S.; Kim, M. G., Lab-on-paper for all-in-one molecular diagnostics (LAMDA) of zika, dengue, and chikungunya virus from human serum. Biosens Bioelectron 2020, 165, 112400.

36. Berendzen, K.; Searle, I.; Ravenscroft, D.; Koncz, C.; Batschauer, A.; Coupland, G.; Somssich, I. E.; Ulker, B., A rapid and versatile combined DNA/RNA extraction protocol and its application to the analysis of a novel DNA marker set polymorphic between Arabidopsis thaliana ecotypes Col-0 and Landsberg erecta. Plant Methods 2005, 1 (1), 4.

37. Anjam, M. S.; Ludwig, Y.; Hochholdinger, F.; Miyaura, C.; Inada, M.; Siddique, S.; Grundler, F. M. W., An improved procedure for isolation of high-quality RNA from nematode-infected Arabidopsis roots through laser capture microdissection. Plant Methods 2016, 12 (1).

38. Vandeventer, P. E.; Mejia, J.; Nadim, A.; Johal, M. S.; Niemz, A., DNA Adsorption to and Elution from Silica Surfaces: Influence of Amino Acid Buffers. The Journal of Physical Chemistry B 2013, 117 (37), 10742–10749.

39. Waggoner, J. J.; Gresh, L.; Mohamed-Hadley, A.; Balmaseda, A.; Soda, K. J.; Abeynayake, J.; Sahoo, M. K.; Liu, Y.; Kuan, G.; Harris, E.; Pinsky, B. A., Characterization of Dengue Virus Infections Among Febrile Children Clinically Diagnosed With a Non-Dengue Illness, Managua, Nicaragua. The Journal of infectious diseases 2017, 215 (12), 1816–1823.

40. Waggoner, J. J.; Gresh, L.; Vargas, M. J.; Ballesteros, G.; Tellez, Y.; Soda, K. J.; Sahoo, M. K.; Nunez, A.; Balmaseda, A.; Harris, E.; Pinsky, B. A., Viremia and Clinical Presentation in Nicaraguan Patients Infected With Zika Virus, Chikungunya Virus, and Dengue Virus. Clinical infectious diseases : an official publication of the Infectious Diseases Society of America 2016, 63 (12), 1584–1590.

41. Vandeventer, P. E.; Mejia, J.; Nadim, A.; Johal, M. S.; Niemz, A., DNA adsorption to and elution from silica surfaces: influence of amino acid buffers. J Phys Chem B 2013, 117 (37), 10742–9.

42. Waggoner, J. J.; Abeynayake, J.; Sahoo, M. K.; Gresh, L.; Tellez, Y.; Gonzalez, K.; Ballesteros, G.; Guo, F. P.; Balmaseda, A.; Karunaratne, K.; Harris, E.; Pinsky, B. A., Comparison of the FDA-approved CDC DENV-1-4 real-time reverse transcription-PCR with a laboratory-developed assay for dengue virus detection and serotyping. Journal of clinical microbiology 2013, 51 (10), 3418–20.

43. Waggoner, J. J.; Abeynayake, J.; Sahoo, M. K.; Gresh, L.; Tellez, Y.; Gonzalez, K.; Ballesteros, G.; Pierro, A. M.; Gaibani, P.; Guo, F. P.; Sambri, V.; Balmaseda, A.; Karunaratne, K.; Harris, E.; Pinsky, B. A., Single-reaction, multiplex, real-time rt-PCR for the detection, quantitation, and serotyping of dengue viruses. PLoS neglected tropical diseases 2013, 7 (4), e2116.

44. Waggoner, J. J.; Gresh, L.; Mohamed-Hadley, A.; Ballesteros, G.; Davila, M. J.; Tellez, Y.; Sahoo, M. K.; Balmaseda, A.; Harris, E.; Pinsky, B. A., Single-Reaction Multiplex Reverse Transcription PCR for Detection of Zika, Chikungunya, and Dengue Viruses. Emerging infectious diseases 2016, 22 (7), 1295–7.

45. Rojas, A.; Stittleburg, V.; Cardozo, F.; Bopp, N.; Cantero, C.; Lopez, S.; Bernal, C.; Mendoza, L.; Aguilar, P.; Pinsky, B. A.; Guillen, Y.; Paez, M.; Waggoner, J. J., Real-time RT-PCR for the detection and quantitation of Oropouche virus. Diagnostic microbiology and infectious disease 2020, 96 (1), 114894.

